# Mapping publication outputs, collaboration networks, research hotspots, and most cited articles in systematic reviews and meta-analyses of medicine and health sciences in Ethiopia: analyses of 20 years of scientific data

**DOI:** 10.1101/2022.02.24.22271416

**Authors:** Tesfa Dejenie Habtewold, Nigussie Tadesse Sharew, Aklilu Endalamaw, Henok Mulugeta, Getenet Dessie, Nigus G. Asefa, Getachew Mulu Kassa, Wubet Alebachew Bayih, Mulugeta Molla Birhanu, Balewgize Sileshi Tegegne, Andreas A. Teferra, Abera Kenay Tura, Sisay Mulugeta Alemu

**Author notes:** Corresponding author: Tesfa Dejenie Habtewold, Ph.D., Department of Quantitative Economics, School of Business and Economics, Maastricht University, Tongersestraat 53, 6211LM, Maastricht, Netherlands.

## Abstract

**Introduction:** Although the publication of systematic reviews (SR) and meta-analyses (MA) has substantially grown in Ethiopia, no robust study systematically characterized these SR and MA was conducted. Thus, we aimed to map publication outputs, collaboration networks, research hotspots, and most cited SR and MA of medicine and health sciences in Ethiopia.

**Methods:** We conducted a bibliometric study of SR and MA published up to December 31, 2021, and systematically searched via PubMed, PsycInfo, EMBASE, and Web of Science databases. We included all SR and MA in medicine and health sciences fields in Ethiopia irrespective of the authors’ affiliation and place of publication. Full records and cited references’ meta-data were extracted from the Web of Science Core Collection database. VOSviewer software was used to perform bibliometric analyses. The relevance of an item (e.g. author, country, or keywords) was measured by its weight based on frequencies using the full or binary counting method) and strength of the link between items was measured using total link strength.

**Results:** In total, 422 SR and MA were published between 2001 and 2021 by 14 research groups (i.e. overall, 1,066 authors participated) who affiliated with institutions from 33 countries. The largest number of SR and MA were published by authors affiliated with Debre Markos University, University of Gondar and Bahir Dar University. In addition, strong collaboration was observed among authors affiliated with institutions in Ethiopia, the Netherlands, Australia, and Canada. The identified research hotspots were maternal and child health, depression and substance use, cardiometabolic diseases, infectious diseases, HIV/AIDS, hepatitis and nutrition. The most cited SR was about domestic violence against women published in 2015. The SR and MA were published in 160 journals, with a majority published in PLOS (11%) and BMC (25%) journals.

**Conclusions:** In this study, we provide a comprehensive summary of collaboration networks, research hotspots, and most cited SR and MA to gain a deeper understanding of the landscape of SR and MA research in Ethiopia. We believe that our study informs researchers, higher institutions, and policymakers about research hotspots and gaps in medicine and health sciences research in Ethiopia. The national and international collaboration is promising, and a concerted effort among researchers, policymakers and funding agencies could increase research outputs and broaden research areas.

## Introduction

Medical knowledge traditionally differs from other domains of human culture by its progressive nature and the need for clear standards or criteria for diagnosing, treating, and identifying clinical improvements and advances.^1^ Health sciences are becoming more evidence-based with the emergence of new methodologies and technologies to meet these standards and to explore the underlying reasons for complex health problems.^1^ One of the major goals of health sciences is to achieve precision in measuring and solving health problems. Clinicians and researchers can get the best up-to-date evidence on a particular topic of interest within a quick exploration of relevant research databases. The overwhelming production of literature with often contradictory and irreplicable findings is the main challenge to get relevant and accurate evidence in medicine and health sciences fields.^2^ In recent years, however, methodologists in medicine and health sciences have become increasingly interested in balancing the accuracy or precision while improving the reliability and generalizability of research findings. Among these, systematic reviews (SR) and meta-analyses (MA) are quickly becoming a popular tools in synthesizing evidence from primary studies.^1^

SR and MA are powerful tools to precisely estimate health service coverage and disease burden, and in recommending effective and efficient interventions for use in daily health care practice. Thus, they are indispensable for the practice of evidence-based medicine and decision-making to guide health policy and practice.^3^ It has often been claimed that the number of SR and MA being published has increased steadily over recent years at the national, regional, and global levels.^4^ In PubMed, 167,029 records were indexed as SR or MA in 2021 (search date December 24, 2021) compared to 22,774 SR or MA indexed in 2017.^4^ In Ethiopia, our research group previously found out that 17 SR and 35 MA were published in medicine and health sciences fields.^5^

Efficient use of SR and MA evidence needs robust estimates of publication trends and characterization. A bibliometric study is a cross-sectional, cross-discipline study, which quantitatively analyzes thousands of publications (i.e., primary studies, reviews, editorials/commentaries, books, and other media communication) as a research object by using mathematical, statistical, and philological methods using text data mining from bibliographic databases.^6,7^ Bibliometric methods use data from citation databases to measure, monitor, visualize and study publication trends, research gaps as well as the impact of scientific outputs.

In Web of Science (search date: January 26, 2022), ‘bibliometric analysis’, ‘bibliometric study’, ‘scientometric analysis’, or ‘scientometric study’ terms were indexed in the title of 6,797 records. In addition, Bibliometric analysis has been used in many research fields including internal medicine, neurosciences/neurology, surgery, psychology, and health care sciences.^8^ Bibliometric analyses are used to examine the global literature outputs on specific topic, such as healthcare ^9^, schizophrenia ^10^, infectious diseases ^11^, asthma ^12^, suicidal behavior.^13^

Given the large publication of SR and MA in Ethiopia during the last few years and their importance in decision-making and guideline development, analysis of research trends and gaps would inform future direction and priority setting for key stakeholders. However, to date, no bibliometric study has been conducted to investigate the growth in publications and the research landscape of SR and MA in medicine and health sciences in Ethiopia. Therefore, we conducted a bibliometric study to systematically investigate and map publication outputs, collaborations, research hotspots and most cited articles in medicine and health sciences fields in Ethiopia using a large sample of SR and MA. This will help researchers, practitioners, and institutions identify research hotspots, emerging trends and remaining gaps in medicine and health sciences research in Ethiopia.

## Methods

### Protocol

The protocol for this study was registered with the Open Science Framework (OSF) (https://osf.io/vapzx). Additionally, all the data used in this manuscript and supplementary materials are available in OSF (https://osf.io/q5dw2/).

### Search strategy

SR and MA in Ethiopia were searched in PubMed (NCBI), PsycInfo (EBSCOhost), EMBASE (direct access), and Web of Science (direct access) international databases from inception until December 31, 2021. “Ethiopia” and “Ethiop*” terms combined by “OR” Boolean operator were searched in the title, abstract and keywords fields. The search was filtered by article, publication, or methodology type (i.e., meta-analysis, review, systematic review, metasynthesis, meta synthesis, meta analysis, literature review) and species (i.e., human). In the Web of Science Core Collection database, given they are not available as filters, the terms “systematic review”, “meta-analy*”, “meta analy*”, and “meta synthesis” combined by ‘‘OR” Boolean operator were separately searched in the title, abstract, keywords and keywords plus. Our database search was supplemented by hand searching of Google, national journals and cross-references to retrieve potentially relevant studies. Our search did not include grey literature and unpublished/preprint SR and MA databases.

### Inclusion and exclusion criteria

SR and MA that fulfilled the following criteria were included. First, the article title must be identified as an SR and/or MA. For articles that were not identified as a SR, MA, or were deemed ambiguous, we reviewed relevant information in the method and result sections, and referred to Cochrane guidelines^14^ to decide the inclusion of the article. Second, irrespective of the place of publication or authors’ affiliation, SR and MA must be based on medicine and health sciences primary studies. Third, the SR and MA must be published in journals indexed in the Web of Science database because the analyses were done using data obtained from this database. Original and updated versions of SR and MA, and title or topic duplicates were considered as separate publications and were included in our analysis as they have different publication dates, separate number of citations, and include different authors and affiliations. However, SR and MA protocols, non-systematic reviews (e.g., scoping, historic, literature, or narrative reviews), exact duplicates (i.e., all the title and authors were the same), conference abstracts, grey literature, commentaries and letters to the editors, reviews following case reports, and SR and MA on animal subjects were excluded. In addition, SR and MA based on international primary studies were excluded. Furthermore, SR and MA without full text were excluded after three attempts to obtain full-text (i.e., contacting corresponding authors, searching on ResearchGate, searching on Sci-Hub).

### Screening, selection, and data extraction

All retrieved records from respective databases were imported to EndNote X9 software^15^ and then to Covidence.^16^ First, duplicates were automatically removed by Covidence and followed by manual removal when automatic removal failed. Then, the title and abstract screening was independently conducted by two reviewers (TD and SM) using Covidence. Full-text of all relevant SR and MA was also independently reviewed by two reviewers (TD and NT) using a priori inclusion criteria. Disagreements during screening and full-text review were resolved by discussion and consultation with a third reviewer. Meta-data including full records and cited references of each SR and MA were extracted from the Web of Science Core Collection database. The full record includes title, authors and their information, source, abstract, all keywords (author keywords and keywords plus), funding, publisher, categories/classification, document information, and journal information. Web of Science provides a common search language, navigation environment, and data structure allowing researchers to search broadly across various resources and use the citation connections inherent to the index to navigate relevant research results and assess their impact. Since the 1900s, the Web of Science Core Collection is a recognized database that indexes >21,894 journals plus books and conference proceedings in natural sciences, health sciences, engineering, computer science, and materials sciences.^17^ It is one of the most commonly used databases for many bibliometric studies.^10-13^ The screening and selection of articles were conducted in accordance with PRISMA guideline.^18^

### Data analyses

All bibliometric text data extracted from the Web of Science Core Collection database were exported to VOSviewer software^19,20^ for bibliometric analyses. We also used ArcGIS to make national and global maps and R software to summarize relevant bibliometric indicators. To evaluate the strength of the link between items, we used Total Link Strength (TLS), which is automatically calculated by VOSviewer upon the mapping of the selected item of analysis. The TLS is proportional to the degree of link, where a higher TLS value indicates a greater relationship between items. Bibliometric indicators were presented as top active ones, most occurrences, or most citations based on item frequency distribution.

For network mapping, the threshold value of weights (i.e., the measure of the relevance of an item) was set as follows: (1) the minimum number of co-authorship was three; (2) the minimum number of institutional and international collaboration was one; (3) the minimum number of occurrences for a keyword and term in the SR and MA was five and 10 respectively; (4) the minimum number of a citation for SR and MA was zero; (5) the minimum number of SR and MA, and citations of a journal for bibliographic coupling analysis was two and zero respectively; and (6) minimum number of co-citation for cited references was seven. Additionally, the minimum cluster size was set to five items per cluster. Structured abstract labels (e.g., introduction or background, objective, methods, results, conclusions), copyright statements, and weakly connected items were excluded during text analyses and visualization of network maps. The full counting method (i.e., each link has the same weight) was used for co-authorship (unit of analysis were authors, organizations, and countries), co-occurrence of keywords (unit of analysis were all keywords), bibliographic coupling (unit of analysis or items were sources/journals), and co-citation (unit of analysis were cited references) analyses. For citation analysis, the unit of analysis was SR and MA. Texts in the title and abstract fields were analyzed to create a term co-occurrence network map and a binary counting method (i.e., only the presence at least once or absence of a term in the SR and MA) was used. Details on the terminologies, types of analyses, counting methods, unit of analysis, and interpreting network visualization maps are presented in the VOSviewer original paper^19^ and its manual.^21^

## Results

### Search results

In total, 4,021 records were retrieved through searching PubMed (n = 1,357), PsycInfo (n = 117), EMBASE (n = 1,174), CINAHL (n = 439) and Web of Science (n = 934) databases. After removing duplicates (n = 1,584), 2,437 articles were screened. Of these, 1,915 records were excluded and 522 SR and MA were selected for full-text review. Five SR and MA were excluded because of the inaccessibility of full-texts after several attempts. After full-text review, 71 regional or international, 15 non-systematic, and three animal studies SR and MA were excluded. Besides, 46 SR and MA were excluded because they were published in journals not indexed in the Web of Science database. Through hand searching, we found an additional 38 records and 14 of them fulfilled our inclusion criteria. Finally, 422 SR and MA were included for bibliometric analysis (Fig. 1).

**Fig. 1:**
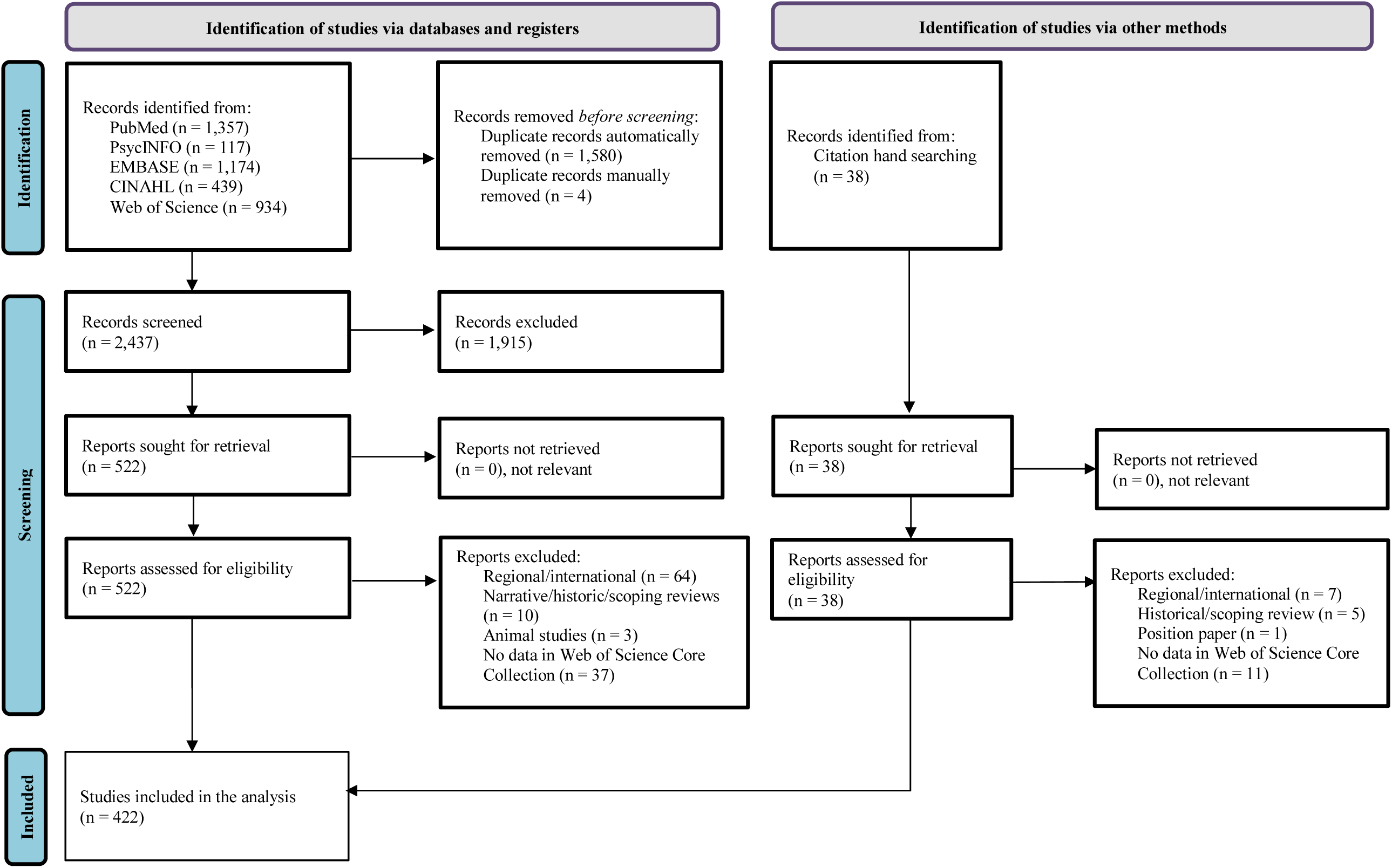
PRISMA flow diagram of identification, screening, and selection process of SR and MA.

### Active authors

In total, 1,066 authors were involved in publishing the 422 SR and MA, which was an average of 2.53 authors per paper. We observed approximately 14 research groups (clusters) were actively working on publishing SR and MA. Of the 1,066 authors, 126 authors who have published at least three SR and MA, and strong collaboration were included in cluster analysis and network visualization mapping (Fig. 2). The to 10 most active authors who published the highest number of SR and MA were Endalamaw A (n = 23), Dessie G (n = 19), Alebel A (n = 19), Kassa GM (n = 18), Habtewold TD (n = 16), Wagnew F (n = 16), Mulugeta H (n = 15), Negesse A (n = 15), Desta M (n = 15) and Demis A (n = 13).

**Fig. 2:**
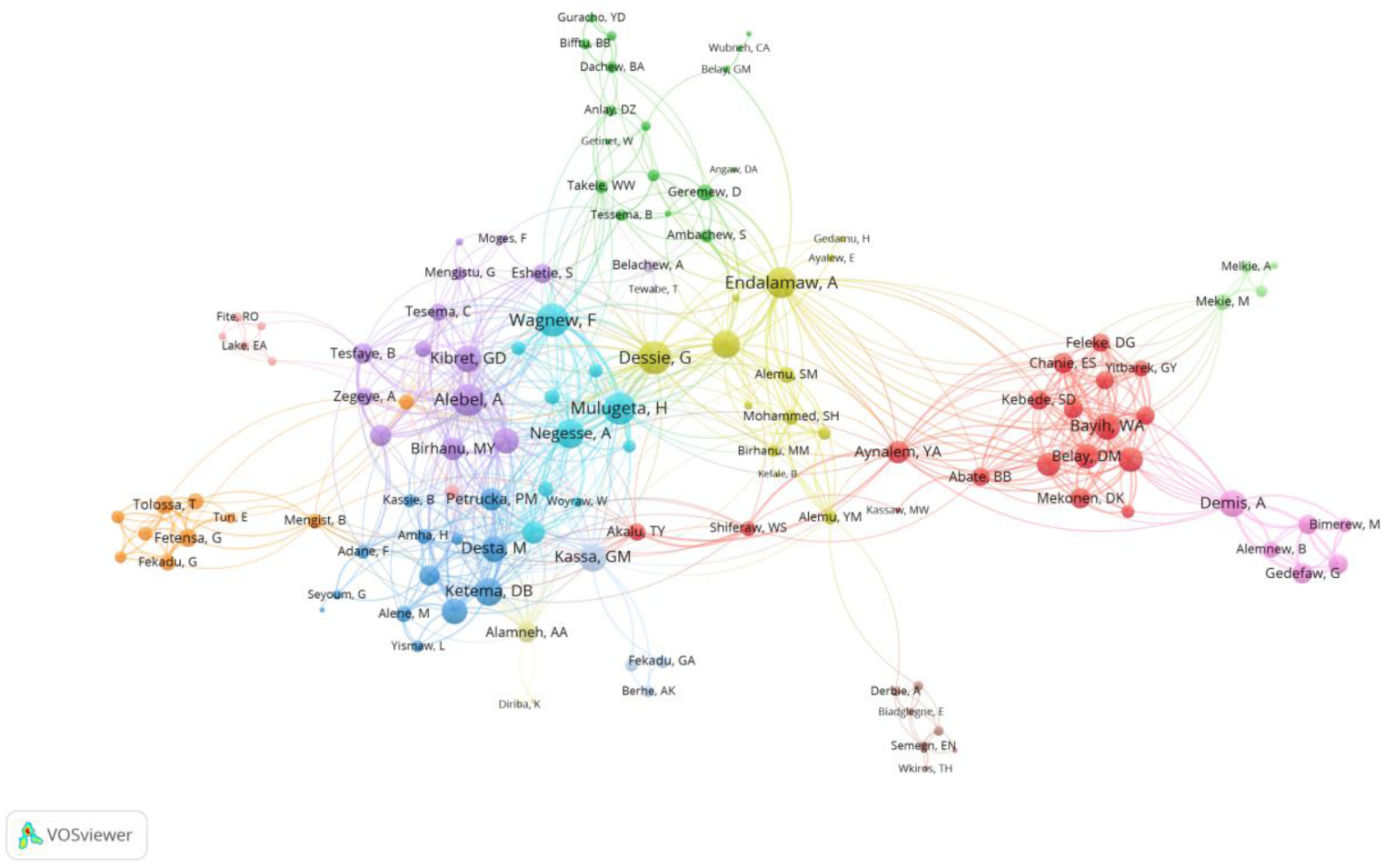
Network visualization map of authors’ collaboration. The node size represents the number of SR and MA published by the author, where the larger node size and label represent the most productive authors. The thickness of the connecting line (aka link strength or edge weight) represents to the strength of collaboration between authors, where the large link strength represents the strong collaboration. The different colors represent different clusters of authors or research group.

### Active institutions

In total, 169 organizations participated in publishing these SR and MA. Fig. 3 shows the network map of 151 organizations that were involved in publishing at least one SR and MA, and with strong collaboration. The top 10 most active organizations that published the highest numbers of SR and MA were Debre Markos University (n = 94), University of Gondar (n = 87), Bahir Dar University (n = 81), Addis Ababa University (n = 49), Debre Tabor University (n = 37), Haramaya University (n = 30), Woldia University (n = 27), Jimma University (n = 25), Hawassa University (n = 21) and Debre Berhan University (n = 18).

**Fig. 3:**
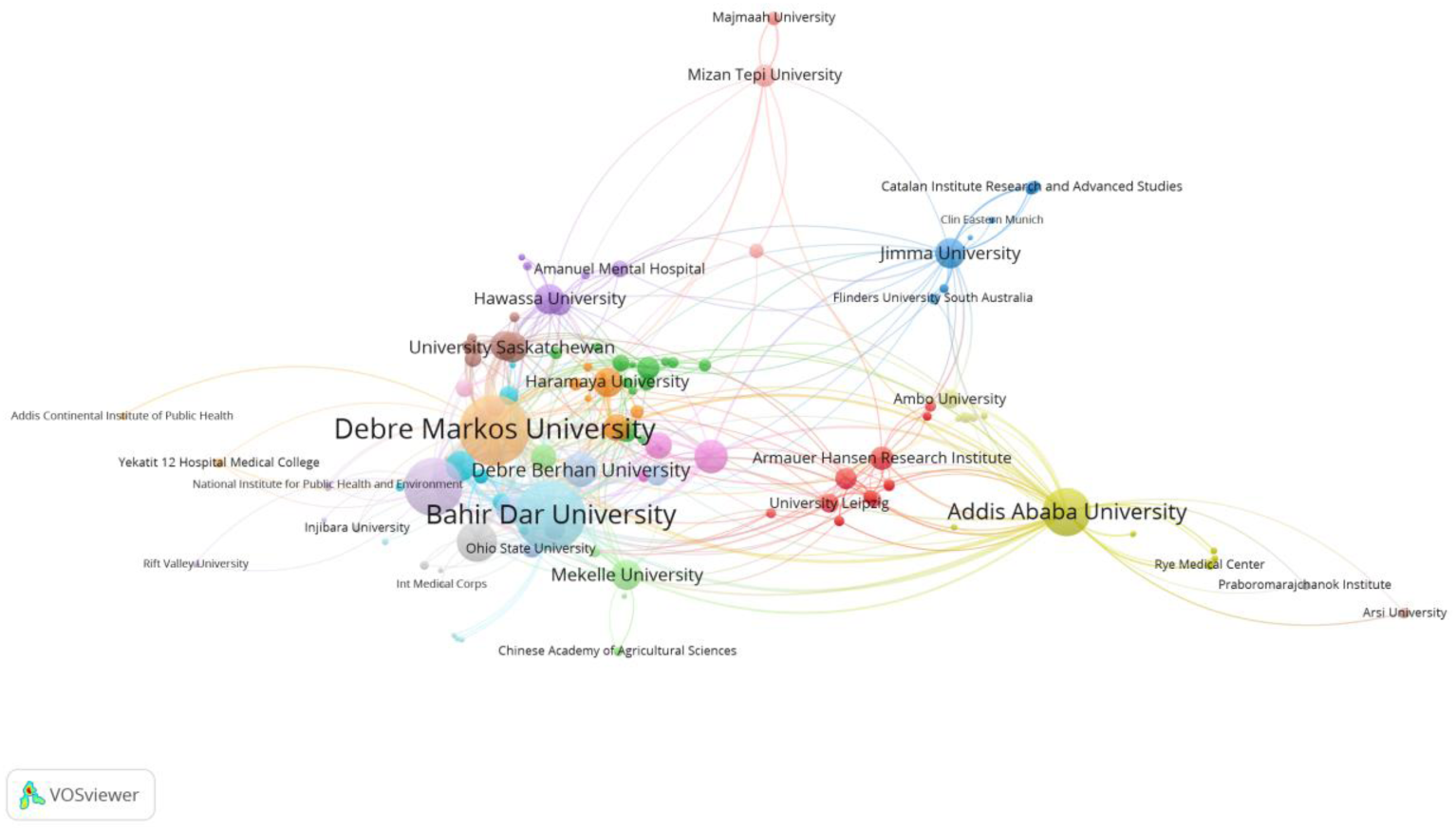
Network visualization map of organizations collaboration. The node size represents the number of SR and MA published by the institution, where the larger node size and label represent the most productive institutions. The thickness of the connecting line (aka link strength or edge weight) represents to the strength of collaboration between institutions, where the large link strength represents the strong collaboration. The different colors represent different clusters of institutions.

### Active regions and countries

Nationally, authors affiliated to institutions located in almost all regions published SR and MA. The largest number of SR and MA, which accounts for the 70.4% (297/422), were published by authors affiliated to institutions in Amhara Region and followed by Oromia Region (24.9%, 105/422) and Addis Ababa City Administration (23.5%, 99/422) (Fig. 4). Globally, authors affiliated with institutions from 33 countries were represented. Most of the SR and MA were published by authors affiliated with Ethiopian institutions (n = 411) followed by Australia (n = 35), The Netherlands (n = 20) and Canada (n = 14) (Fig. S1). As shown in Fig. S2, the strongest collaboration was observed between Ethiopia and Australia (total link strength = 32) followed by Ethiopia and The Netherlands (total link strength = 20), and Ethiopia and Canada (total link strength = 14).

**Fig. 4:**
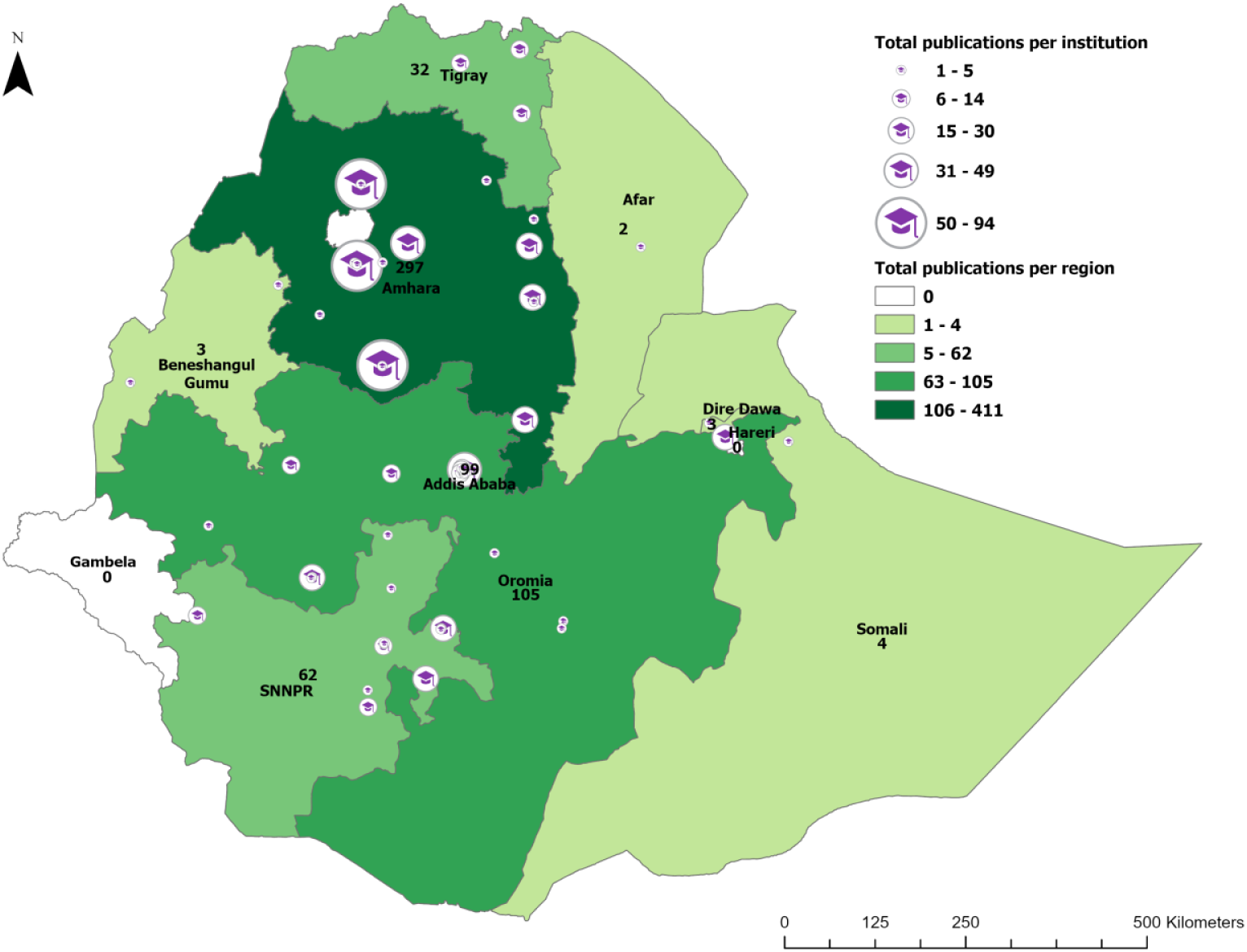
Map of institutional and regional SR and MA publication outputs. The icons represent location of the institutions, and the size represents the total number publications of the institution.

### Research hotspots

In total, 1,233 keywords were used and 132 of them were used in at least five SR and MA, and included in cluster analysis and network visualization mapping. Overall, we identified seven research hotspots (Fig. 5). *Cluster one* (red) contained 29 keywords mainly focusing on maternal and child health. *Cluster two* (green) contained 26 keywords mainly focusing on infectious diseases. *Cluster three* (blue) contained 20 keywords mainly focusing on depression and substance use. *Cluster four* (yellow) nutrition contained 18 keywords mainly focusing on nutrition. *Cluster five* (purple) contained 16 keywords mainly focusing on HIV/AIDS. *Cluster six* (light blue) 13 keywords mainly focusing on hepatitis. *Cluster seven* (orange) contained 10 keywords mainly focusing on cardiovascular diseases. The top 10 most frequently used keywords were Ethiopia, determinant, meta-analysis, prevalence, systematic review, women, children, HIV, risk, district, health, epidemiology, Addis Ababa, university and mortality (Fig. 6).

**Fig. 5:**
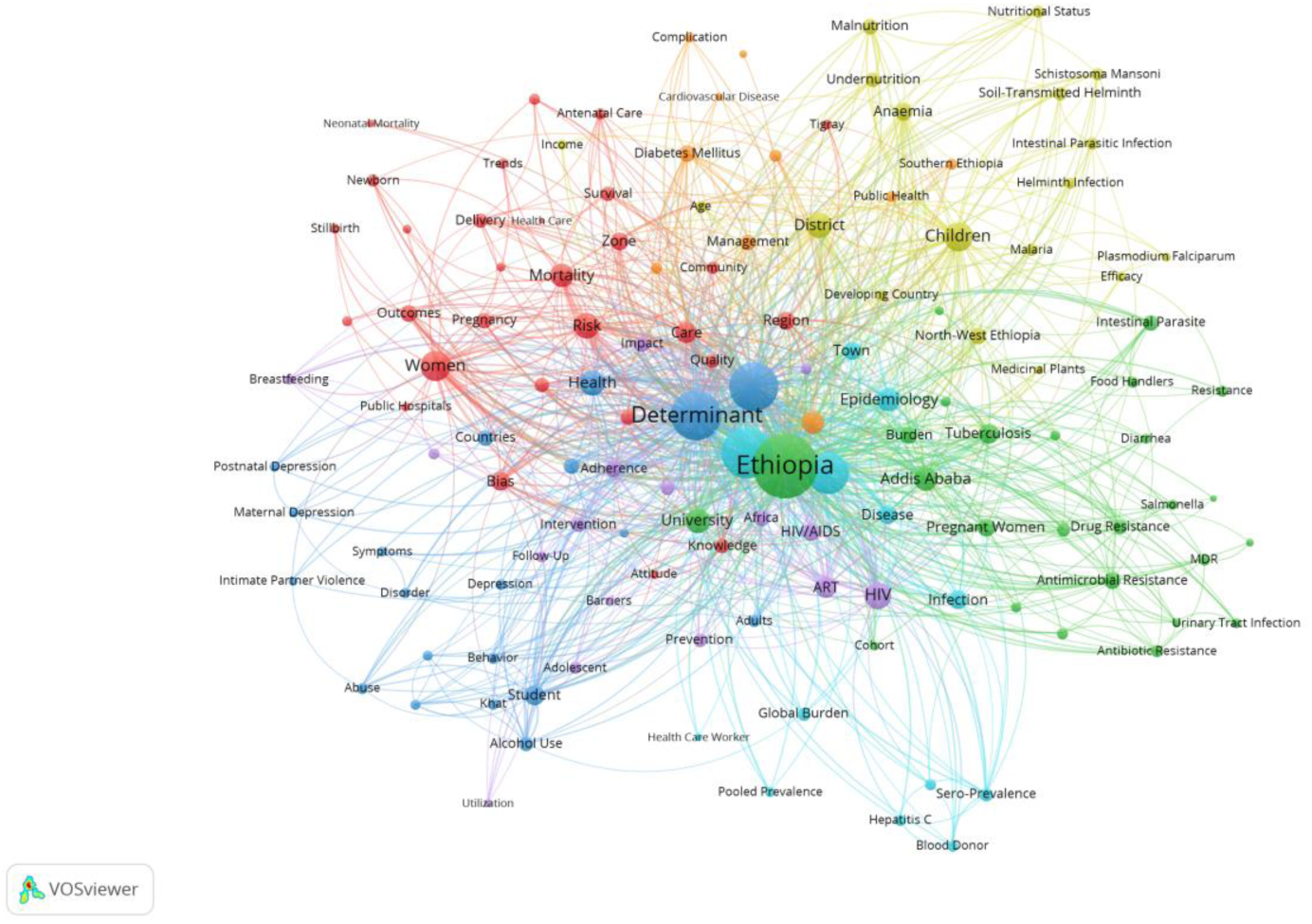
Network visualization map of keywords and research hotspots in SR and MA. The node size represents the frequency of the occurrence the keywords, where the large node size and label represent the most frequently used keywords. The thickness of the connecting line (aka link strength or edge weight) represents to the frequency of co-occurrence between keywords, where the large link strength represents the most frequent co-occurrence. The different colors represent different clusters of keywords or research hotspots.

**Fig. 6:**
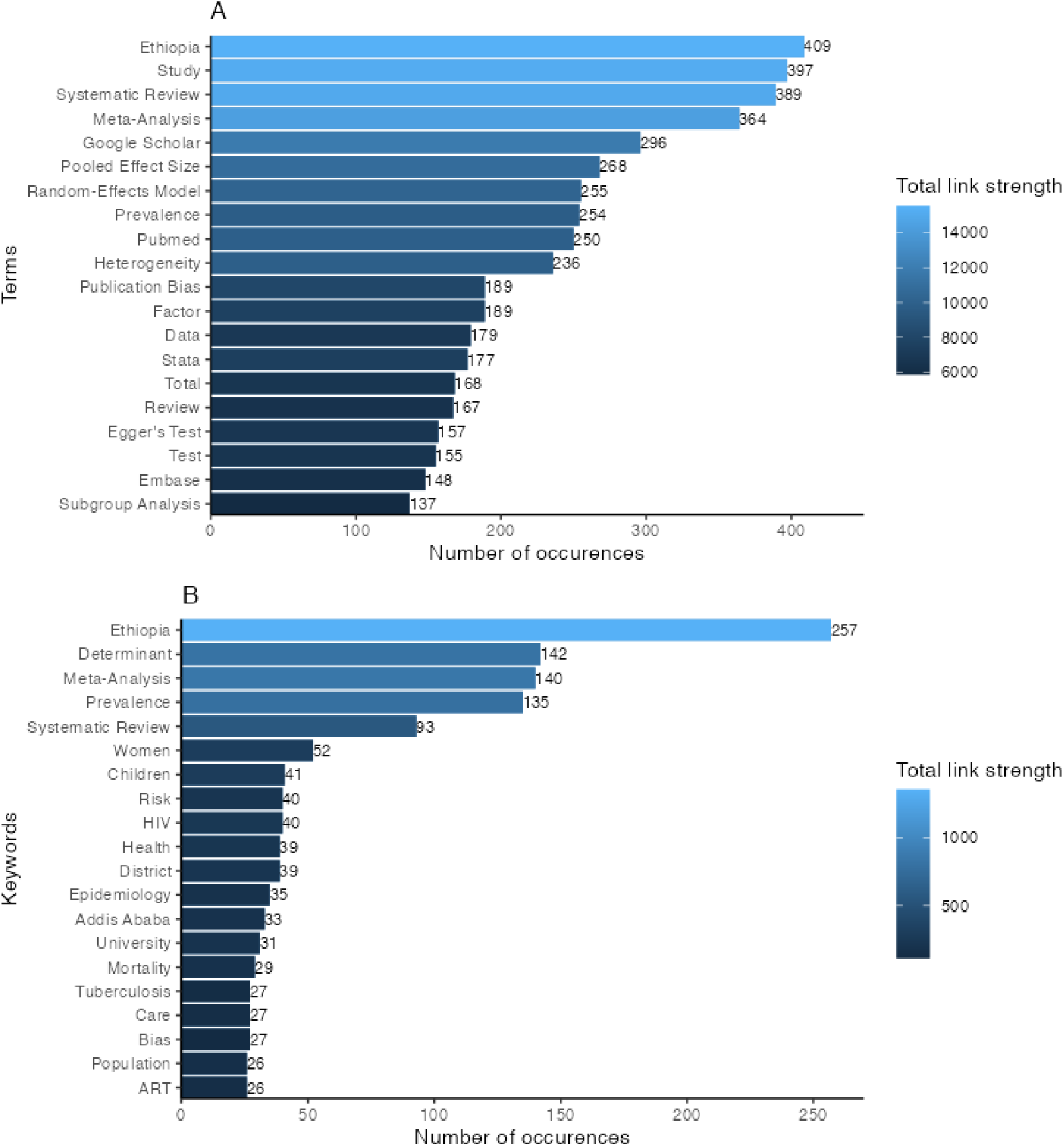
Top 20 most frequently indexed terms (Panel A) and keywords (Panel B).

### Terms co-occurrence

There were 6,459 terms used in the title and abstract fields and 331 terms that occurred in at least 10 SR and MA were included in the term co-occurrence analysis and network visualization map (Fig. S3). The top 10 most frequently used terms in the title and abstract fields were Ethiopia, study, systematic review, meta-analysis, Google scholar, pooled effect size, random-effects model, prevalence, PubMed, heterogeneity, publication bias, determinant, data and STATA (Fig. 6).

### Active journals

The SR and MA were published in 160 different journals, of which 57 journals that published in at least two SR and MA were presented in the network visualization map (Fig. 7). The top 10 most active journals were PLOS ONE (46 SR and MA, 268 citations), BMC Infectious Diseases (27 SR and MA, 287 citations), BMC Public Health (19 SR and MA, 124 citations), Reproductive Health (14 SR and MA, 176 citations), BMC Pregnancy and Childbirth (14 SR and MA, 86 citation), Biomed Research International (12 SR and MA, 24 citations), Systematic Reviews (10 SR and MA, 33 citations), Archives of Public Health (9 SR and MA, 20 citations), Heliyon (9 SR and MA, one citation), Italian Journal of Pediatrics (6 SR and MA, 53 citations), BMC Pharmacology and Toxicology (6 SR and MA, 44 citations) and BMJ Open (6 SR and MA, 39 citations). In total, the top 10 journals published 42.2% (178/422) of SR and MA and received 1,155 citations that accounted for 52.8% (1,155/2,187) of the total citations. Of note, all of these journals were open access.

**Fig. 7:**
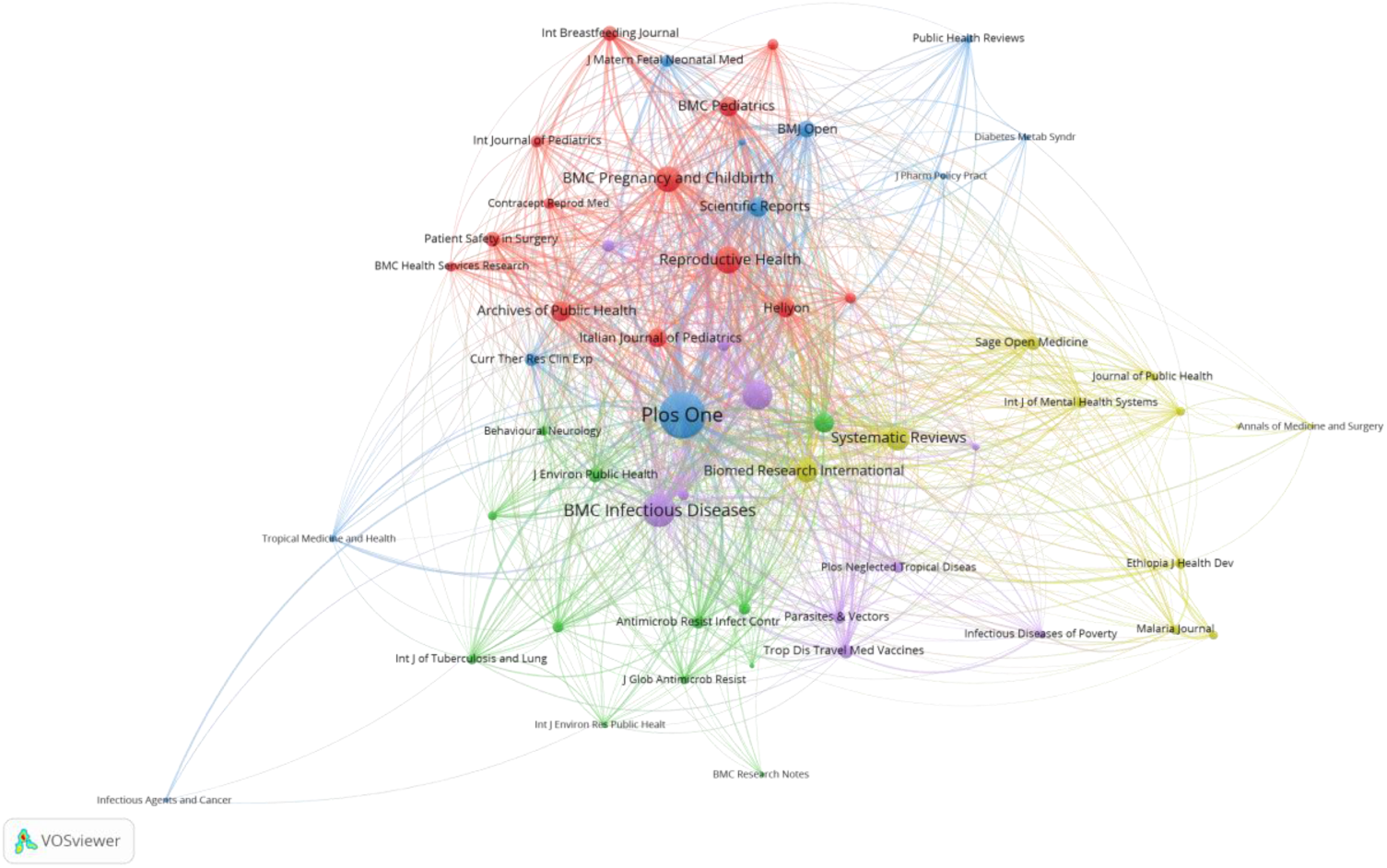
Network visualization map of journals that published the SR and MA. The node size represents the number of cited references in the SR and MA from the journal, where the large node size and label represent the most cited journal by the SR and MA. The thickness of the connecting line (aka link strength or edge weight) represents to the frequency of shared cited reference, where the large link strength represents the large number cited reference from these journals. The different colors represent different clusters of journals.

### Most cited articles

In this analysis, all the 422 SR and MA were included for density visualization mapping (Fig. S4). As shown in Table 1, the top 10 most most cited SR and MA are Semahegn A and Mengistie B (74 citations)^22^, Misganaw A et al (67)^23^, Belyhun Y et al (54)^24^, Kibret KT and Mesfin YM (53)^25^, Ayalew MB (41)^26^, Kebede A et al (40)^27^, Alebel A et al (39)^28^, Tesfaye G et al (36)^29^, Eshetie S et al (34)^30^, and Abdulahi A et al^31^ and Berhe AK et al (32 each).^32^

**Table 1:**
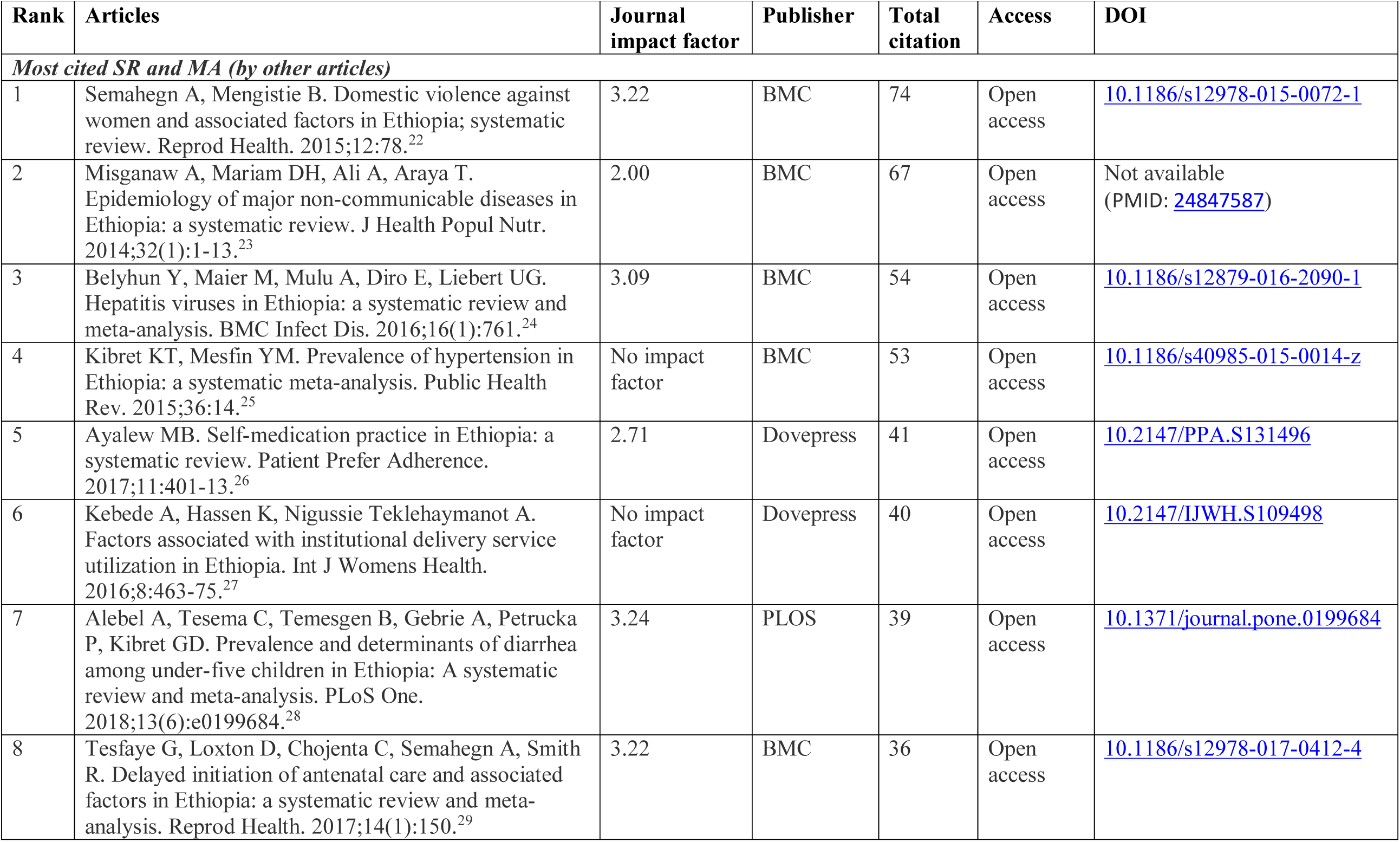
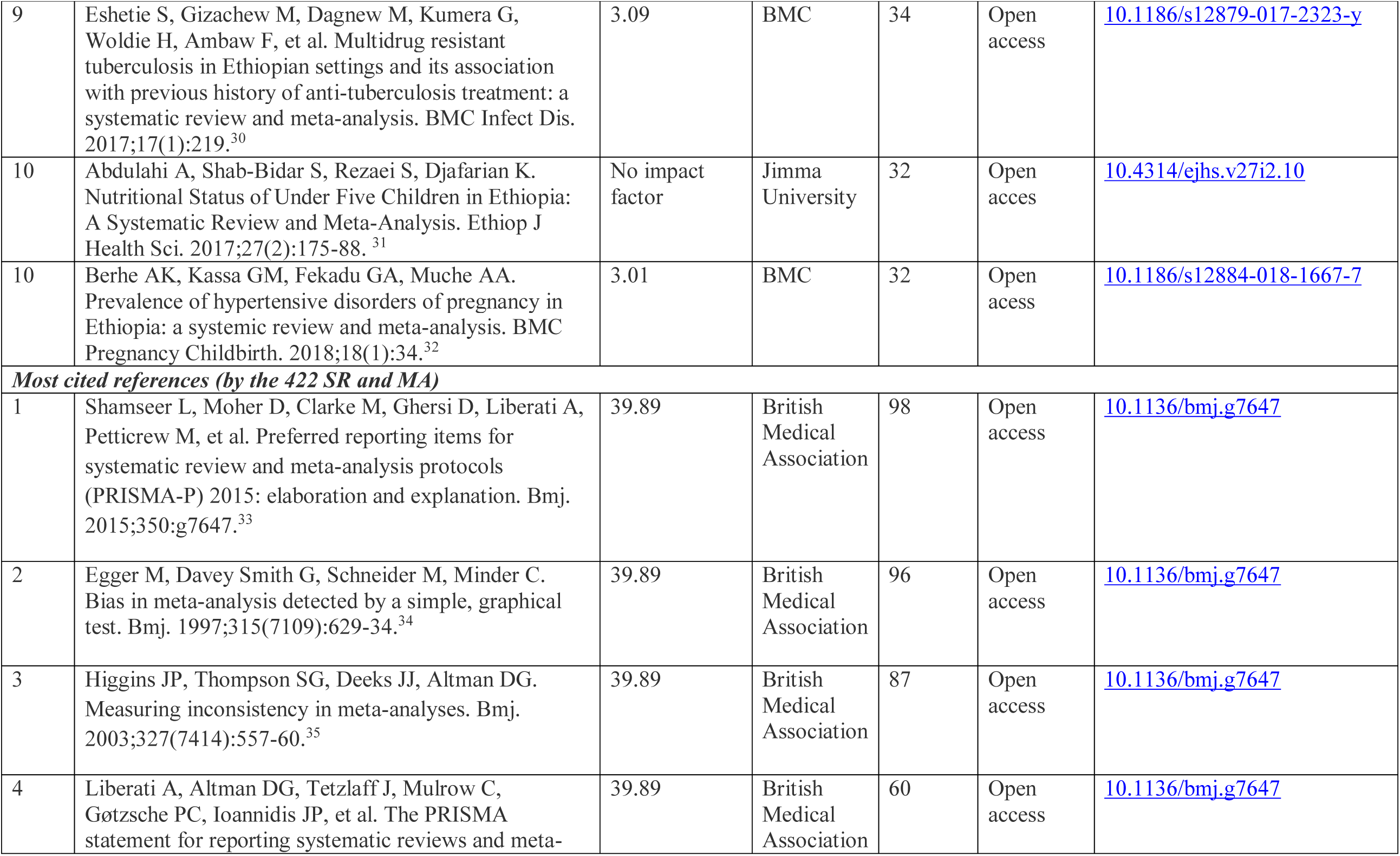
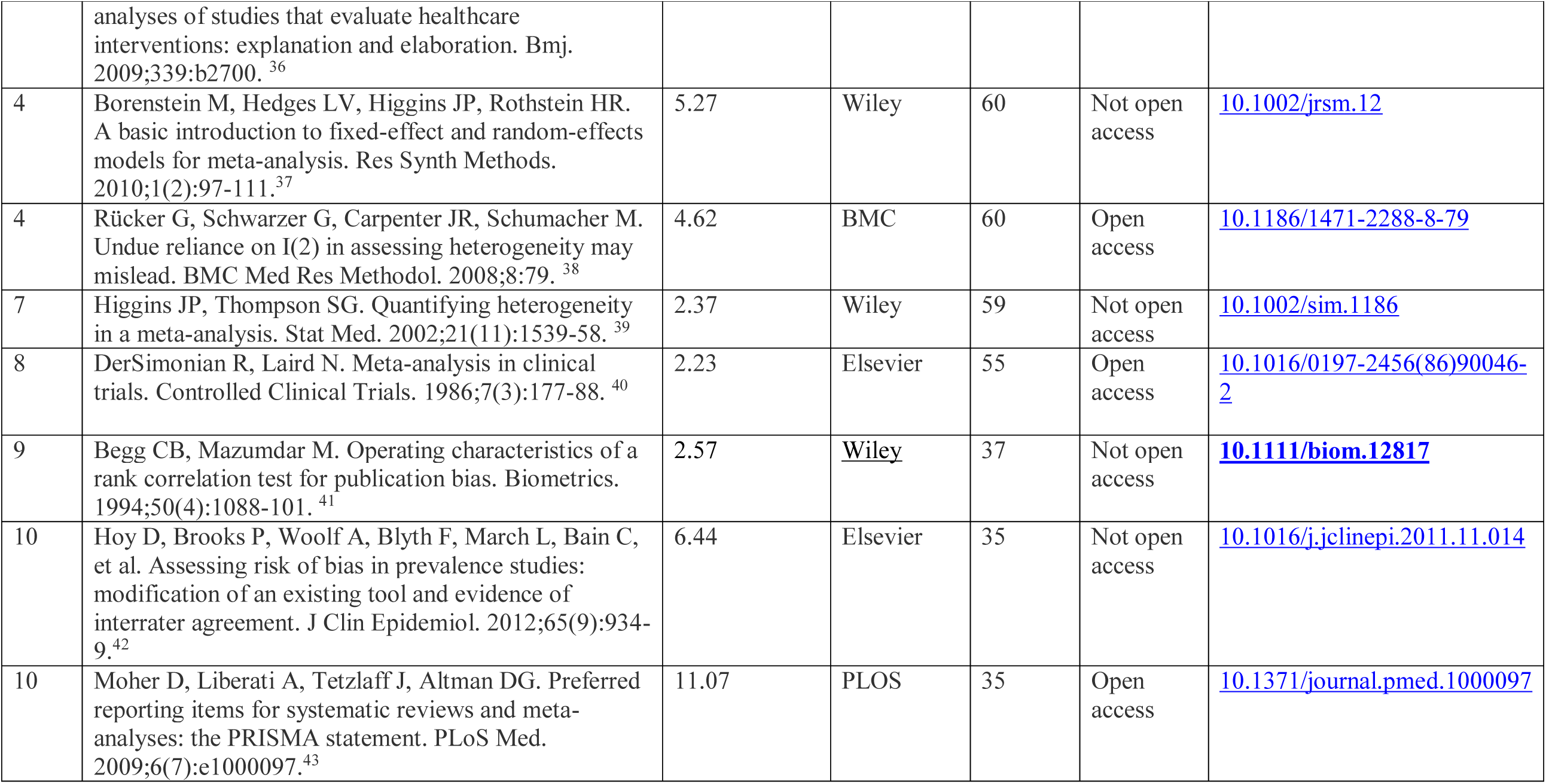
Top 10 most cited SR and MA, and references ranked by total citation up to December 31, 2021.

The co-citation analysis showed that 19,362 references were cited by the 422 SR and MA, which was an average of 45.56 references per SR and MA. Of these, 71 references were cited by at least seven SR and MA, and presented in the network visualization map of 71 frequently cited references (Fig. S5). The most cited references were Shamseer I (98 citations)^33^, Egger M (96)^34^, Higgins JPT (87)^35^, Liberati A (60)^36^, Borenstein M (60)^37^, Rucker G (60)^38^, Higgns JPT (59)^39^, Dersimonian R (55)^40^, Begg CB (37)^41^, Hoy D (35)^42^, Moher D (35)^43^, Huedo-medina TB (30)^44^, Sterne JAC (28)^45^, Munn Z (23)^46^, Nyaga VN (22)^47^, and Stang A (20).^48^ Details of these most cited articles are shown in Table 1.

## Discussion

To the best of our knowledge, this is the first comprehensive bibliometric study of SR and MA of medicine and health sciences in Ethiopia. In total, 1,066 authors affiliated with 169 institutions that spanned across 33 countries were involved in publishing the 422 SR and MA in 160 different journals. Through analyzing authors’ keywords and keywords generated by Web of Science, we identified seven research hotspots in medicine and health sciences. The most frequently used keywords and terms were Ethiopia, determinant, meta-analysis, prevalence and systematic review. We also found that SR and MA were more frequently published by authors affiliated with Debre Markos, Bahir Dar and Gondar universities. Moreover, we observed that the most most cited SR was conducted on domestic violence against women and the most cited reference was on PRISMA guideline.

The network of authorship in this study revealed collaborations among authors from different institution and countries indicating that SR and MA are a type of publication, which can facilitate the exchange of scientific knowledge and promote collaboration among researchers.^9^ Thus, future authors who plan to conduct SR and MA studies can communicate and collaborate with these authors and institutions to share experience and further advance the implementation of SR and MA methods. Besides, a strong and sustainable national and international collaboration among authors in different countries is needed to increase the publication rate, maintain its trend and ensure the quality of SR and MA studies. These collaborations can also promote a better research culture that supports all individuals involved and may help overcome contextual (e.g., social, economic, cultural environment) and methodological (e.g., specialized skills) challenges to conduct SR and MA. Institutional collaborations maybe more effective than individually motivated collaboration for involving authors from multiple disciplines. Generally, the publication rate and trend of SR and MA in Ethiopia is very promising, but also far behind in terms of quantity and quality, and in its infancy stage compared to the output in developed countries.^5^ Besides, the topics addressed by the SR and MA are skewed and most active authors are affiliated to limited number of institutions.

All of the top publishing journals are fully open access and have an impact factor from 2.48 to 3.41 (Q2 and Q3 rank). The fact that they are open access implies it is more likely that authors from Ethiopia and other developing countries freely access articles and more frequently cited by researchers. In addition, BMC and PLOS journal offers waivers for article processing charges (APCs) for papers whose corresponding authors are based in low-income countries, which remarkably helped Ethiopian authors to overcome APC challenges. Authors who want to publish SR and MA in medicine and health sciences may consider these journals as a priority.

The top 10 keywords that appeared in at least five SR and MA were ‘Ethiopia’, ‘determinant’, ‘meta-analysis’, ‘prevalence’, ‘systematic review’, ‘women’, ‘children’, ‘HIV’, ‘risk’ and ‘district’. The fact that frequent occurrence of these keywords can be interpreted as a sign of the nature of SR and MA that most of them were focused on the prevalence and associated factor studies. Overall, we observed that maternal and child health, depression and substance use, cardiometabolic diseases, infectious diseases, HIV/AIDS, hepatitis and nutrition are research hotspots in medicine and health sciences. This fact may be a result of both the research interests of the scientific community and the number of existing primary studies in medicine and health sciences fields. Additionally, this may be due to the high burden of communicable diseases, maternal, childhood and nutritional conditions, neglected tropical diseases, non-communicable diseases, injuries, and public health emergencies, which makes them a priory area for research and intervention.^49^ On one hand, our finding shows that the burden of these problems is currently high and future studies, preventive measures and curative intervention strategies may target these hotspot research areas. On the other hand, the result implies that future studies may focus on other research topics to minimize duplication of work and broaden the discovery of science. For example, there were fewer publications in the fields of ophthalmology, anesthesiology, otorhinolaryngology, dermatology, health economics, and medical imaging. Research activity and funding should be directed towards less explored healthcare topics and address new research questions.^9^ Our result is based on the most frequently used keywords in published SR and MA, and it does not mean other research areas were not entirely investigated. Generally, keywords cluster analysis can facilitate researchers to see research hotspots and understand the research direction on the field.

The most cited SR published by Semahegn A and Mengistie B was ‘Domestic violence against women and associated factors in Ethiopia; systematic review’.^22^ The results of this review indicated that lifetime prevalence of domestic violence against women by husband or intimate partner ranged from 20% to 78 % and was significantly associated with alcohol consumption, chat chewing, family history of violence, occupation, educational status, residence, and decision making power.^22^ The other frequently cited SR and MA were conducted on non-communicable diseases ^23,25,32^, diarrhea ^28^ and nutritional status ^31^, institutional service utilization ^27^ and initiation of antenatal care ^29^, self-medication ^26^ and hepatitis virus ^24^, and MDR-TB.^30^ This is also in line with the research hotspots identified. This may be because all were published in fully open access journals. Additionally, this may be due to the increasing global burden of non-communicable diseases, tuberculosis, and hepatitis that leads to design different types of studies and developing guidelines. We believe these SR and MA could have an important impact on the subject and future authors may be frequently cited and become most cited if they will publish SR and MA on these topic areas.

The most cited reference published by Shamseer was ‘Preferred reporting items for systematic review and meta-analysis protocols (PRISMA-P) 2015’.^33^ Overall, the most cited references were on PRISMA and other reporting guidelines^33,36,43,46^, meta-analysis model^37,40^, heterogeneity assessment^35,38,39,44^, publication bias assessment^34,41,45^, quality appraisal^42,48^ and statistical analysis software.^47^ In fact, it is not surprising to see these articles are most frequently cited because we included SR and MA that follow similar methodological and analytical procedures. Future researchers can easily focus on these articles to read and design their SR and MA. They are also globally known and used by all researchers who want to conduct SR and MA studies in medicine and health science fields.

In one hand, most medical practice in low- and middle-income countries such as Ethiopia is not evidence-based related to limited resources and poor access to up-to-date information. As a result, most health care decision-making tools are based on evidence from developed countries without considering the cultural and socioeconomic differences, and translation into context was limited. On the other hand, despite having ample evidence on the common problems in the country, the efforts in using the available evidence – for example use and translation of implementation research or action research is poor and the quest for knowledge search is trending without using what exist at hand. For example, a recent study in Ethiopia showed that half of health care providers were unable to find resources for implementing an evidence-based practice that attributed to lack of training and poor health facility infrastructures, such as computers and internet.^50^ Therefore, there is a pressing need to develop an evidence base that addresses the requirements of the developing countries and getting this evidence into the hands of those who deliver healthcare is fundamental to improvements in healthcare delivery and health outcomes.^51^ The findings from this study can provide useful up-to-date evidence on the landscape of SR and MA research and it is a wake-up call to balance generation of new knowledge and implementation by decision-makers.

We believe that the main strength of our study is that fact that we included all SR and MA to observe collaboration networks, research hotspots and most cited SR and MA. In addition, we used well-established and organized bibliometric data from the Web of Science Core Collection, which enabled us to systematically synthesize current state of knowledge. We also believe that our study has some limitations that should be considered by readers. First, data from grey literature and publications in journals not indexed in the Web of Science were not included. This could underestimate the number of SR and MA. However, including journals indexed in the Web of Science, which is also promoted by the Ethiopian Ministry of Education and other agencies, is essential for generating strong and reliable evidence. Second, we did not assess the methodological or reporting quality of included SR and MA since our aim was quantitative analysis of the bibliometric data. Third, authors affiliations might be counted twice since authors with two different country affiliations were counted once for each country. This may increase the research output of certain countries with greater international collaboration even if the authors from that country were not the main or corresponding authors.^11^ Finally, the citation analysis did not take into consideration self-citations, which could create a bias in the number of citations for documents, countries, journals, and authors.

The value of bibliometric studies has been increasing because it provides the opportunity to summarize emerging study designs such as SR and MA. Our research could be further complemented by more detailed analyses of the areas of increased primary studies activity, but with less research activity in SR and MA and vice versa, explore the methods and content for sub-groups of studies (e.g., different types of SR and MA, different healthcare subject categories) and assess the reporting quality of these SR and MA. Future research could provide clarity and descriptive insight into the different types and characteristics of SR and MA. The findings of our bibliometric analysis have the potential to inform key stakeholders (authors, scholarly journals, policymakers, funding agencies) about trends and gaps in the production of SR and MA, and guide future research agenda. As a result, this information may help to avoid duplication of research efforts and wastage of resources.

## Conclusion

The publication of SR and MA in medicine and health sciences has recently seen a significant increase in Ethiopia. A total of 1,066 authors working in 169 institutions that spanned across 33 countries were participated in publishing 422 SR and MA in 160 different journals. We also found that SR and MA were more frequently published by authors affiliated with Debre Markos, Bahir Dar and Gondar universities. Moreover, we identified seven research hotspots in medicine and health sciences: maternal and child health, depression and substance use, cardiometabolic diseases, infectious diseases, HIV/AIDS, hepatitis and nutrition. Our findings give insight that national and international research collaboration is not adequate, whereby most SR and MA publications are clustered in a limited number of authors, institutions, and countries. Thus, national collaborations between SR and MA authors from institutions with increased research productivity and institutions with less research activity should be encouraged to promote the production of SR and MA in areas of healthcare that are less explored. We believe that our study provides researchers, higher institutions, and healthcare policymakers with evidence on the research landscape of SR and MA in medicine and health sciences to update their knowledge and formulate new investigation areas. Our study emphasized the importance of well-planned and organized national and international collaborations and funding to increase the publication rate of primary studies, and SR and MA, broaden research areas and minimize duplication of work.

## Data Availability

All data produced are available online at https://osf.io/q5dw2/.

## Notes

### Competing Interest Statement

The authors have declared no competing interest.

### Clinical Protocols

https://osf.io/vapzx

### Funding Statement

This study did not receive any funding

### Summary of Updates

1. The whole manuscript has been copyedited and revised to make it more clear and precise. 2. Figure 3 revised - abreviation organizational names used in the figure written in full. 3. Authors affiliations updated.

