## Supplementary figures for "Mapping publication outputs, collaboration networks, research hotspots, and most cited articles in systematic reviews and meta-analyses of medicine and health sciences in Ethiopia: analyses of 20 years of scientific data"

**Supplementary results**

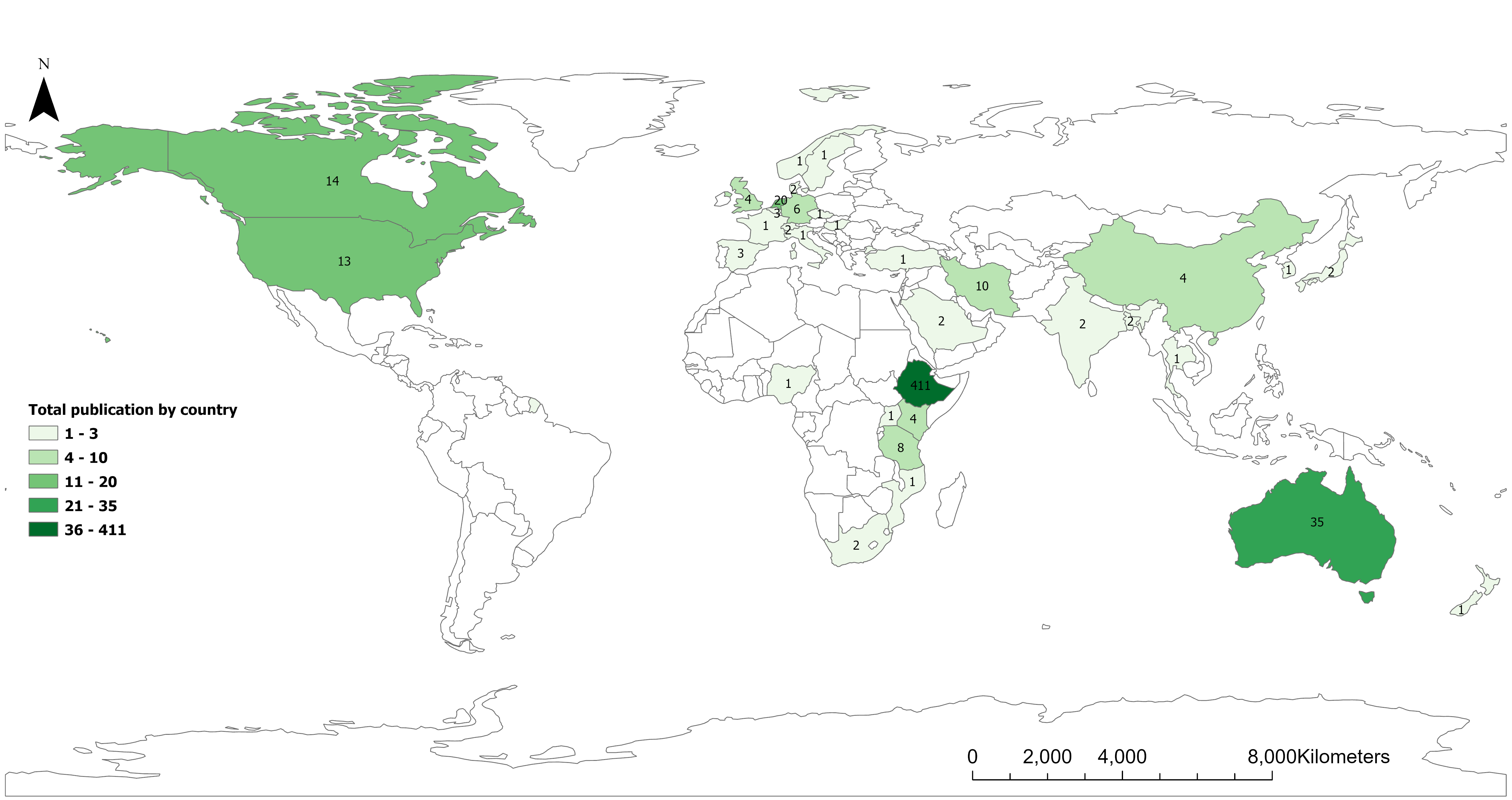

Fig. S1: Countries publishing SR and MA in Ethiopia.

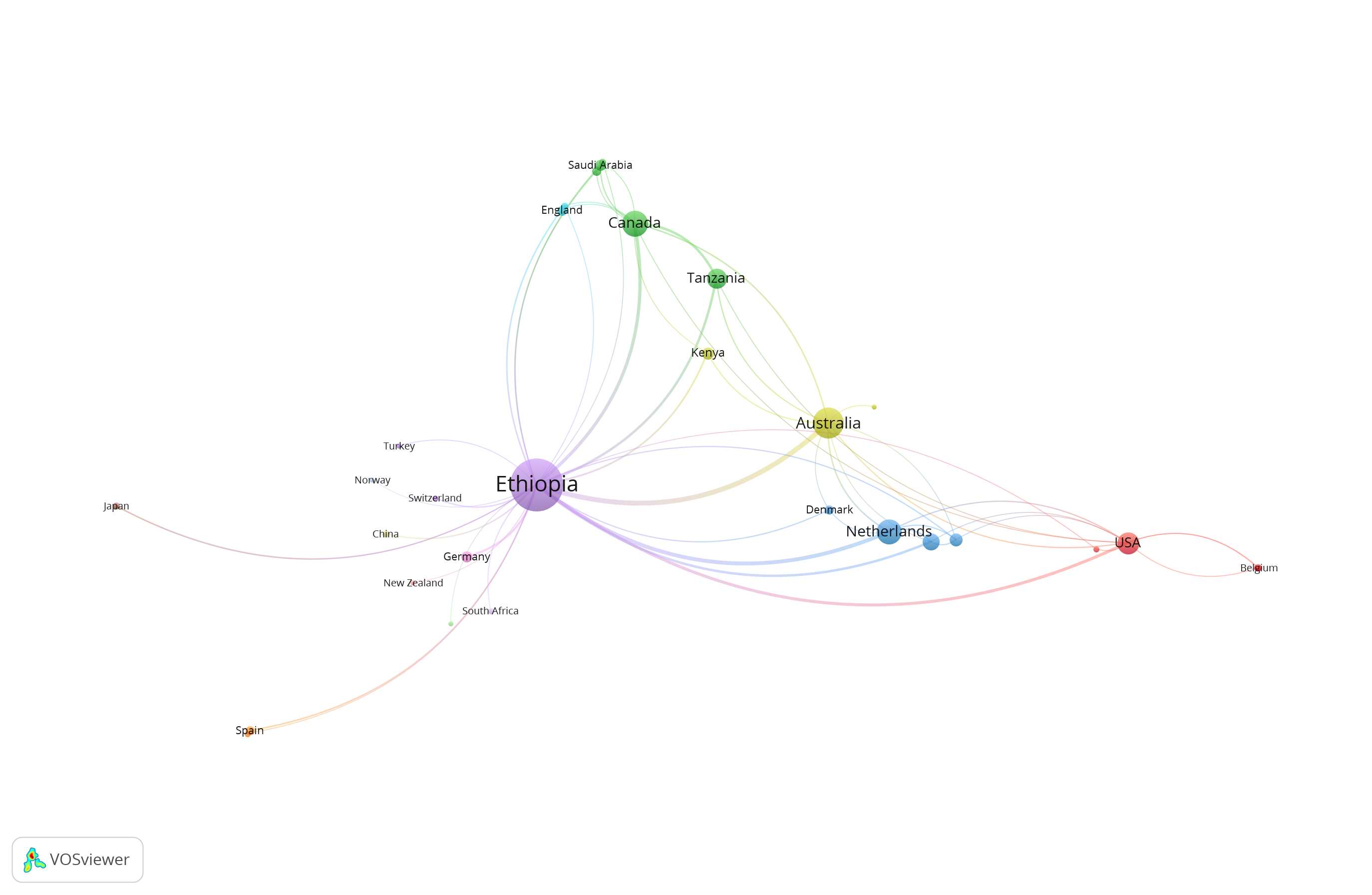

Fig. S2: Network visualization map of international research collaboration.

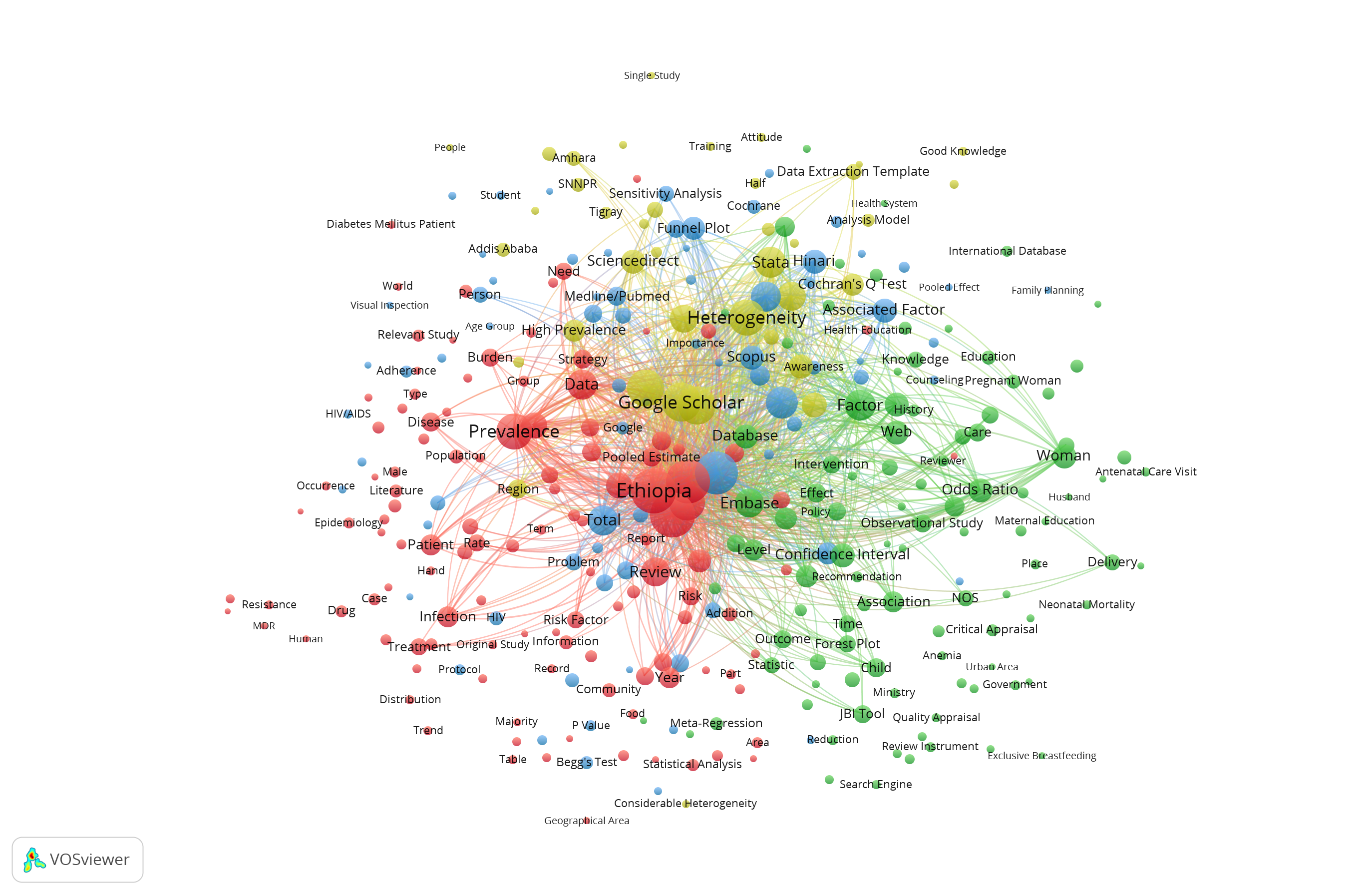

Fig. S3: Network visualization map of terms co-occurrence in the title and abstract fields.

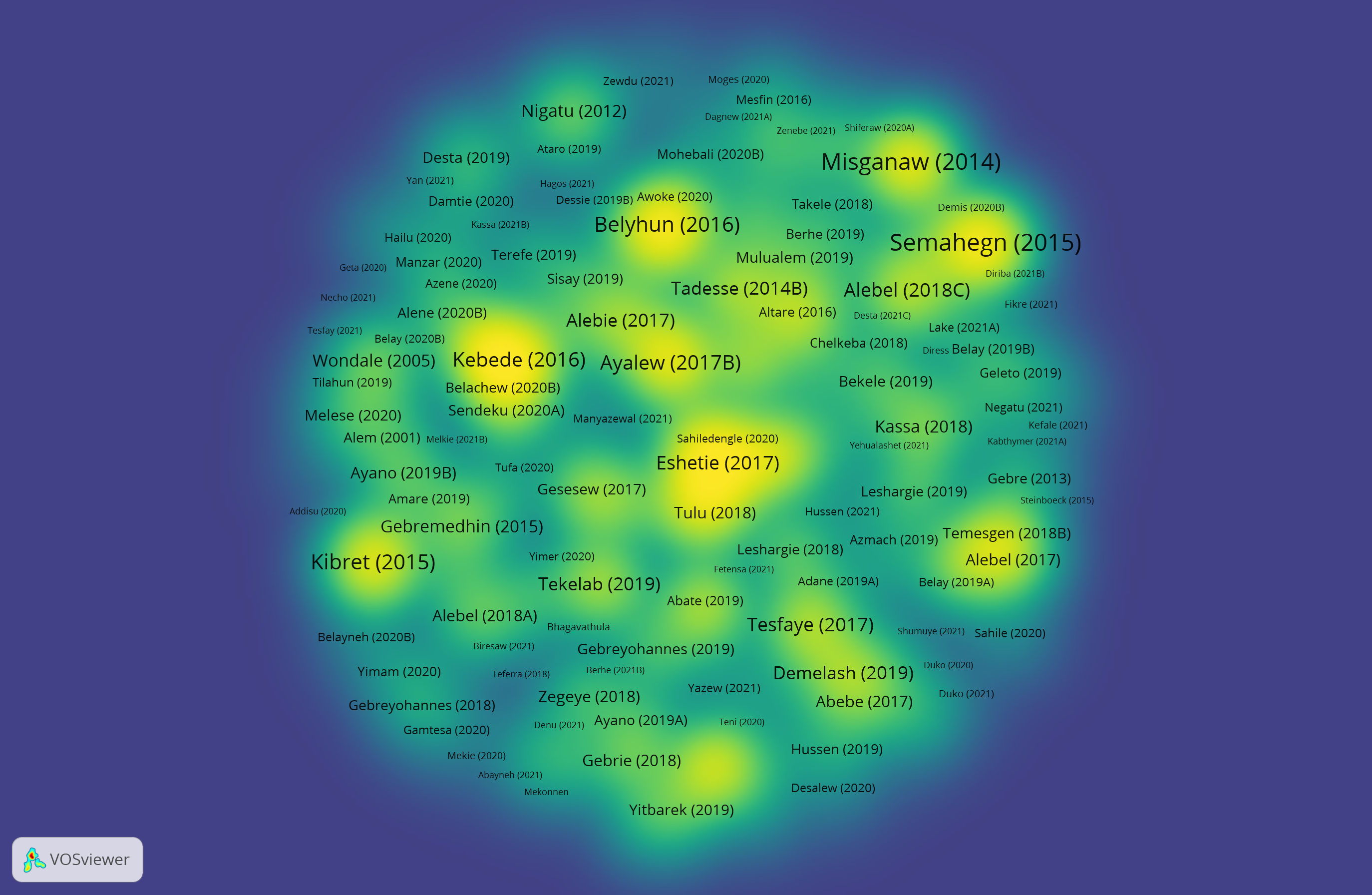

Fig. S4: Density visualization citation map of the 422 SRs and MA. The color yellow represents the most cited SR and MA and the color blue represents the least cited SRs and MA.

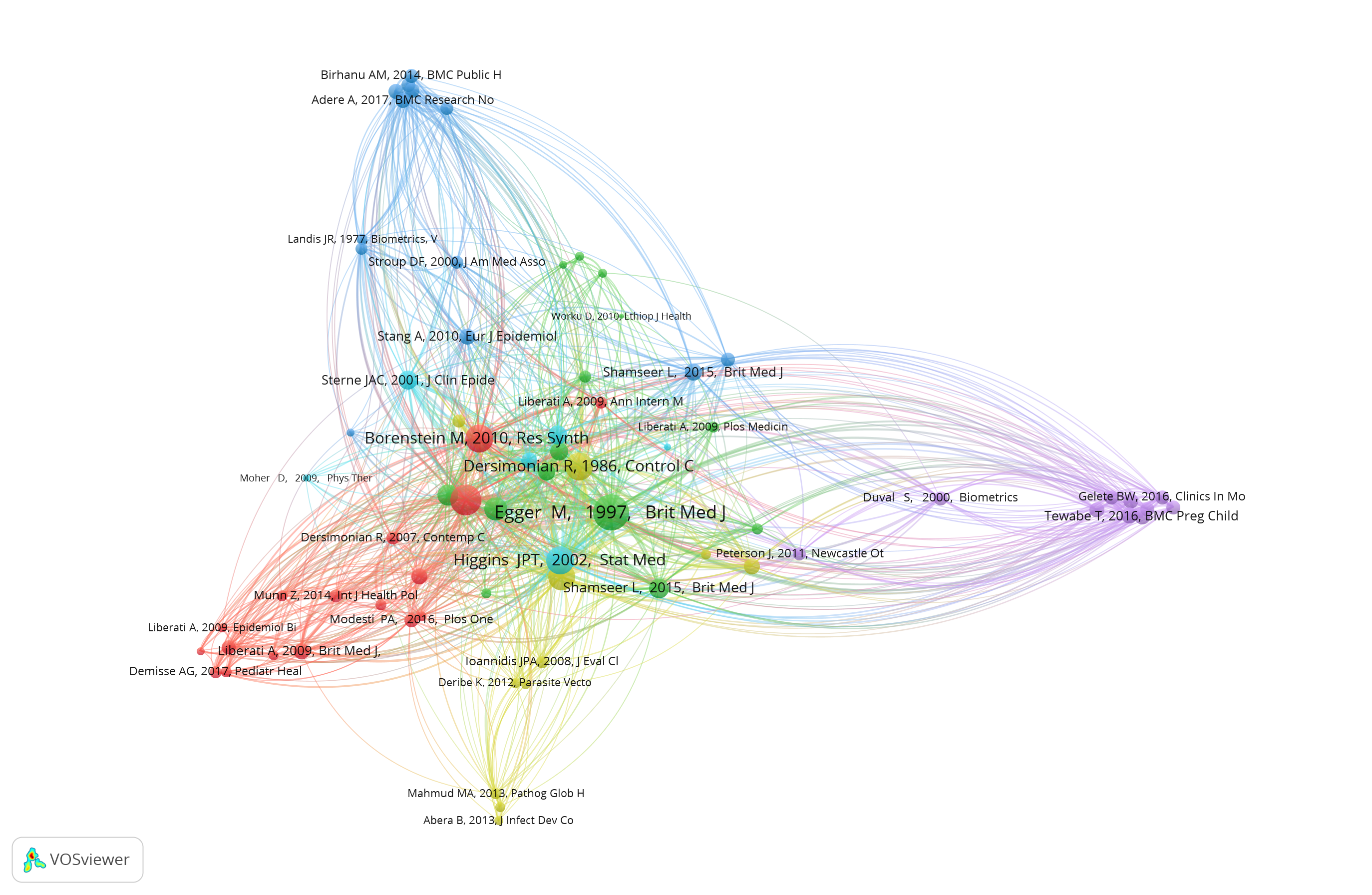

Fig. S5: Co-citation network visualization map.
